# Individualized Mechanistic Modeling Reveals Viral Infectivity and CD8^+^ T cell Expansion as Drivers of Heterogeneous Influenza Dynamics

**DOI:** 10.1101/2025.10.24.25338576

**Authors:** Nicole Bruce, Jordan J.A. Weaver, Cailan Jeynes-Smith, Fatemeh Beigmohammadi, Solène Hegarty-Cremer, Amanda P. Smith, Morgan Craig, Amber M. Smith

**Author notes:** Corresponding author: Amber M Smith.

## Abstract

Influenza virus infections vary widely in severity, with human challenge studies revealing substantial heterogeneity in viral shedding and immune responses. However, the mechanistic basis of this variability remains unresolved. Here, we applied individualized mechanistic modeling to determine how variation in viral replication and CD8^+^ T cell responses shape infection kinetics and symptom dynamics once variation in exposure dose is removed in individuals experimentally challenged with H1N1 influenza virus. Our analysis identified four distinct infection clusters driven predominantly by individualized differences in viral infectivity and CD8^+^ T cell expansion rates rather than baseline T cell levels or killing efficacy. These mechanistic distinctions were conserved across independent cohorts and enabled accurate prediction of symptom trajectories despite subjectivity in symptom reporting and differences in viral strain. Collectively, these findings demonstrate that interindividual variation can converge on similar infection trajectories and clinical outcomes and provide a mechanistic basis for improving individual-level prediction of influenza infection dynamics.

## Introduction

Influenza virus infects millions of people each year, and the predictability of developing symptomatic infection is difficult. In addition, there is significant variability in viral shedding patterns, immune responses, and symptom presentation, with viral loads typically being a poor predictor of severity. Cellular immunity, humoral immunity, and cytokines have all been noted as statistical correlates in the risk of symptomatic influenza and in outcome^1–23^, but these do not necessarily lead to strong differences in infection trajectories and may not always be causal. Recent studies have identified substantial variability in baseline immunity due to genetics, environmental exposures, comorbidities, and infection history^3,19,24–32^, which can influence viral replication and the ensuing immune response. However, small differences in viral loads can be accompanied by large differences in cellular immunity^15,33–36^, making the reliable prediction of infection outcome from immune data difficult even when paired with advanced statistical approaches. This highlights the nonlinearity of immunity and the need to account for multivariate interindividual heterogeneity using mechanistic computational frameworks^37^.

Cell-mediated immunity, which is central to the resolution of influenza infections, has been shown significant variability between individuals, where elevated CD4^+^ and CD8^+^ T cells have been correlated to protection from symptomatic infection^3,4,7,11,13,16,17,20,21,23^. However, higher influenza-specific T cell counts or frequencies in the blood are not typically associated with reduced viral shedding and less is known about whether individuals with naturally higher baseline cell counts would be more protected from exposure to a novel influenza virus, such as during a pandemic. Human challenge studies provide a unique perspective on influenza infections in individuals lacking antibody-mediated immunity^18,20,38–53^. These studies control for the timing of infection, virus strain, and dose to facilitate the identification of interindividual variability in viral loads, immune responses, and symptoms due to other intrinsic factors. The data demonstrate that viral shedding, immunity, and symptoms vary widely between individuals, even in cases of mild infection, and that prior infection does not always confer protection. What drives the heterogeneity in infection dynamics remains unclear.

The complexity of immune responses, influenza virus infection, and human biological variation complicates efforts to identify the determinants that shape individual host-pathogen trajectories and outcomes. Thus, we used an individualized mechanistic mathematical modeling approach applied to the largest longitudinally sampled cohort of influenza-infected individuals to identify the sources of variation in viral shedding and CD8^+^ T cell responses while accounting for differential viral dosing, viral replication, baseline CD8^+^ T cell populations, and CD8^+^ T cell functionality. Our analyses revealed that interindividual heterogeneity is driven primarily by variation in viral infectivity and CD8^+^ T cell expansion rates, rather than by baseline T cell levels or cytotoxic efficacy alone. Personalized model realizations further demonstrated that variability in shared infection mechanisms can unite to create non-unique infection trajectories without strong dependence on baseline immunity, and that symptom dynamics are predictable despite their subjective nature and limited correspondence with systemic innate immunity. Collectively, these results emphasize that individualism does not guarantee individualized outcomes and demonstrate how pairing quantitative clinical data with individualized mechanistic modeling enhances the interpretability and predictability of influenza virus infections.

## Materials and Methods

### Influenza Infection, Immune, and Symptoms Following Challenge with H1N1

#### Cohort 1

We used data from the largest study of adult human volunteers challenged with influenza that included longitudinal sampling of host immunity^38^. All participants were healthy adults (aged 18-45 y) confirmed to be seronegative to the inoculating virus prior to enrollment. The volunteers were intranasally infected with 3.5 × 10^6^ TCID_50_ (250 µl; ID numbers beginning with 1), 7 × 10^6^ TCID_50_ (250 µl; ID numbers beginning with 2), or 7 × 10^6^ TCID_50_ (500 µl; ID numbers beginning with 3) influenza A/California/04/2009 (H1N1). Nasal washes were collected daily from days 1 to 7 to quantify viral loads (copies/ml), and blood was collected from -1 to 7 to quantify CD8^+^ T cell frequencies. Of the 35 individuals in the study, 12 had more than one day of positive viral shedding (IDs 103, 107, 110, 111, 112, 204, 207, 302, 307, 308, 311, and 312; denoted here as ‘Cohort 1’). Low viral measurements that occurred between two higher viral load measurements were considered a failed sample and excluded from our analysis (denoted with dashed outlined markers). The reported viral loads for some individuals (IDs 103, 110, 111, 112, 204, and 207) on days 1 and/or 2 post-infection were reported as zero despite confirmed infection. Because it is unclear if these are failed tests, measurements below a limit of detection, or true zero measurements, we imputed the values using the geometric mean of the individuals who had robust measurements (i.e., 6.6 × 10^3^ copies/ml for day 1 and 1.7 × 10^5^ copies/ml for day 2; denoted as open markers). The limitations of data collection made this a preferable approach to have sufficient data for the mathematical model framework.

The matched CD8^+^ T cell frequencies for activated (CD38^+^) effector (CD45RA^+^CD27^-^CCR7^-^) and memory (CD45RA^-^CD27^+^) phenotypes were used because they most closely resemble the dynamics of antigen-specific effector and memory CD8^+^ T cells in the lung from animal studies^34^. These PBMC-derived measurements provide a systemic readout of circulating CD8^+^ T cell dynamics but may not directly capture local mucosal immune responses in the upper respiratory tract. The reported abundances are fractional measurements, expressed as proportions of total PBMCs, rather than absolute cell counts. Because total PBMC counts for each participant were not provided in the original dataset, absolute CD8^+^ T cell numbers were approximated by scaling the abundances by a constant 2.6 × 10^7^ cells for each phenotype per individual. This was rationalized by comparing cell abundances with cell counts from murine data^34^, which suggested relatively similar timescales of CD8^+^ T cell expansion and peaks at the time when CD8^+^ T cells are most active in clearing infected cells (**Figure S1**). However, it is a notable limitation of the data, where total cell counts may differ between individuals.

Total symptom scores were reported by the individuals on a 0-51 scale comprised of 0-3 ratings (0: absent; 1: mild; 2: moderate; 3: severe) for respiratory symptoms (breathing difficulty, cough, hoarseness, nasal stuffiness/congestion, runny nose, sneezing, sore throat, and wheezy chest) and pain (abdominal pain, diarrhea, earache, facial or eye pain/tenderness, fatigue, fever, headache, musculoskeletal ache, and nausea/vomiting).

#### Cohort 2

We used data from a study of 28 adult human volunteers (ages 18-40 y) who were intranasally infected with 1 × 10^5^ TCID_50_ influenza A/Texas/36/91 (H1N1) and exhibited more than one day of viral shedding^39^ (denoted as ‘Cohort 2’). All participants were confirmed to be seronegative to the inoculating virus prior to enrollment. Daily viral loads (TCID_50_) were quantified from nasal washes, and symptom scores were recorded by the infected individuals. Total symptom scores were reported on a 0-36 scale comprised of 0-3 ratings for breathing difficulty, fever, headache, hoarseness, fatigue, coughing, sneezing, sore throat, runny nose, earache, muscle ache, and chest pain.

### Mathematical Model of Primary Influenza Infection

To describe the dynamics of influenza infection, we modified our previously validated mathematical model that describes the interactions between the virus and effector and memory CD8^+^ T cells^34^. Briefly, healthy epithelial cells (*T*) become infected at rate *βV* per day and undergo an eclipse phase (*I*_1_). The eclipse cells transition to productively infected cells (*I*_2_) at rate *k* per day. These cells produce virus (*V*) at rate *p* per cell per day, which is then cleared at rate *c*per day. The infected cells (*I*_2_) are cleared by innate mechanisms at rate *δ*per day and by effector CD8^+^ T cells (_E_ and _EEM_) at a density-dependent rate of δ_E_(I_2_) modeled using a saturating functional response with maximal rate δ_E_ infected cells/T cell/day and half-saturation constant 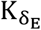 infected cells. Reactivation of memory CD8^+^ T cells (_EIM_) into effector memory cells (_EEM_) occurs at a nonlinear rate of σ(I_1_), modeled as a functional response with maximal rate σper day and half-saturation constant *K*_σ_T cells, and with decay at rate *d*_*R*_ per day. We model the effector CD8^+^ T cell expansion phase using a linear chain trick, where activation of earlier effector cells (_E1_) occurs at rate ξ per infected cell/day. The transition rate through each differentiation state (_Ei+1…n_) occurs at a= n*/*τ_E_ per day, where *n* is the number of states and τ_E_ is the time delay. The terminally differentiated effector CD8^+^ T cells (_E_) transition into influenza-specific memory CD8^+^ T cells (_EM_) at rate ζper day after τ_M_ days. The model is given by Equations (1)-(10).

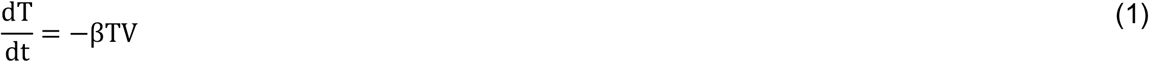

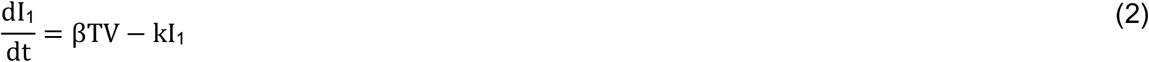

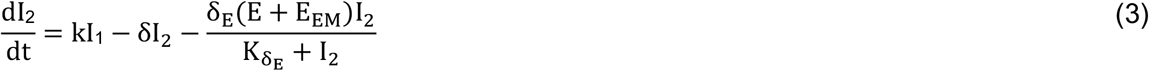

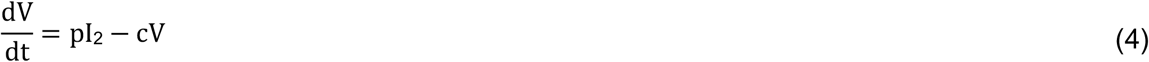

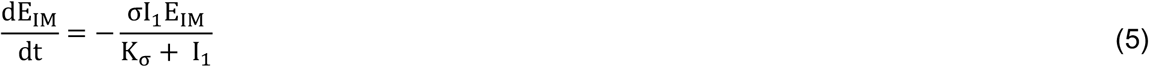

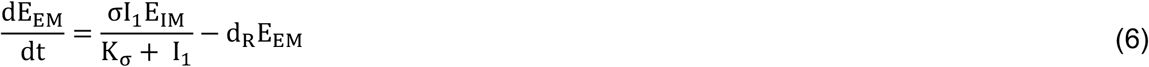

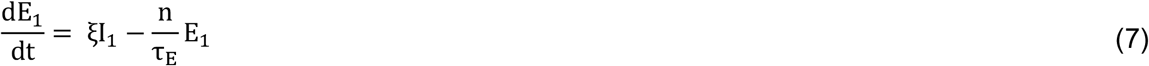

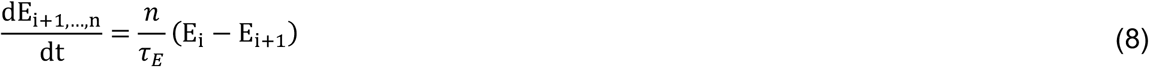

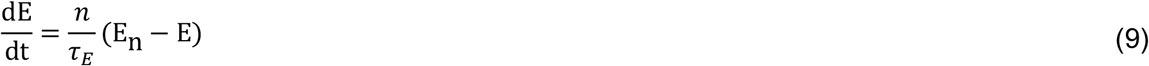

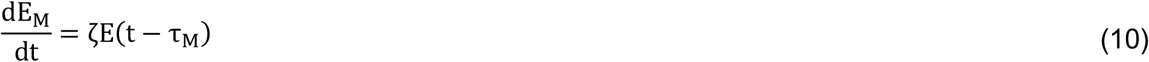

### Parameter Quantification

To quantify the model parameters, the model was fit simultaneously to the viral loads and effector and memory CD8^+^ T cells using a simulated annealing global optimization algorithm with 95% confidence intervals for the population obtained by bootstrapping as previously described^34,54^. Parameter uncertainties for each participant were determined by selecting parameter sets that resulted in a cost within 10% of the best-fit cost from a Monte Carlo simulation. Model simulations were implemented in MATLAB.

Estimated parameters in the primary infection model for both the population and individuals included the rates of viral infectivity (*β*(copies/ml)^-1^ day^-1^); viral clearance (c day^-1^); non-specific infected cell clearance (δ day^-1^); effector CD8^+^ T cell-mediated infected cell clearance (δ_E_ infected cell (T cell)^-1^ day^-1^; 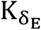 cells); effector memory CD8^+^ T cell reactivation (σday^-1^; K cells) and decay (d_-_ day^-1^); effector CD8^+^ T cell activation (ξ (infected cell)^-1^ day^-1^), number of differentiation compartments (n), and time delay (τ_E_ days); and memory CD8^+^ T cell generation (ζday^-1^) and time delay (τ_M_ days). The eclipse phase transition rate was set to k= 6 per day based on prior studies^34,54^.

The initial number of healthy epithelial cells (T(0)) was set to 1 × 10^7^ cells for the population but fit for the individuals to estimate differences in infection sizes resulting from different doses. This value also represents the infection size due to all target cells becoming infected within the model. The viral production rate (p(copies/ml)/cells/day) was fit for the population and fixed for each individual. The initial number of eclipse cells (I_1_(0)) was set to 75 cells, and the initial free virus (v(0)) and productively infected cells (I_2_(0)) were set to 0^34,54^. The initial effector and memory CD8^+^ T cells (_E_(0) and _EM_(0), respectively) were set to 0 with the baseline values added to the model solution as in our previous studies^34^. The baseline number of influenza-specific memory CD8^+^ T cells (_EMI_(0)) was assumed to be the number formed during the recorded infection (i.e., value at day 7 – value at day 0).

### Clustering Analysis

To cluster individuals by their clinically measured data for viral loads and effector CD8^+^ T cells alone or matched, we used hierarchical clustering^55^ (*scipy*.*cluster*.*hierarchy*.*linkage*) with Ward’s method^56^ in Python. To place the data on the same scale for each individual’s paired data, the data was normalized using *scikit-learn*.*standardscaler*. Optimal clustering was determined using the maximum silhouette score^57^ from *scikit-learn* and the elbow plot method from the within-cluster sum of squares.

To cluster individuals with similar host-pathogen interactions, we performed a principal component analysis (PCA) in Python (*scikit-learn*.*decompose*.*pca*) on the personalized model parameters. To place parameters on the same scale, parameters were log_10_-transformed and then normalized using *scikit-learn*.*standardscaler*. Optimal clustering was determined using hierarchical clustering with Ward’s method, the maximum silhouette score, and the elbow plot method.

### Modeling Symptom Scores

Our prior work showed that the cumulative area under the curve (CAUC) of infected cells (*I*_1_+*I*_2_) accurately predicts the area of infected tissue, which was correlated to symptoms^34,58^. To estimate the symptom scores recorded by each volunteer, the CAUC was calculated using *cumtrapz* in MATLAB, with their symptom resolution modeled by a linear decay from the symptom peak^34^.

## Results

### Interindividual Variation in Viral Shedding and CD8^+^ T cells Patterns

Viral loads in 12 human volunteers who experienced viral shedding for >1 day after being challenged with influenza A/California/04/2009 (H1N1) (Cohort 1) had viral peaks ranging from 5.6 × 10^4^ to 2.1 × 10^7^ copies/ml, with a population average peak of 2.7 × 10^5^ copies/ml on day 2 post-infection^38^ (**Figure 1A**). The population and most individuals’ viral shedding followed a biphasic pattern, with a phase of slower clearance followed by a phase of faster clearance, similar to viral shedding patterns observed in murine primary infection models^34,54,58^. Activated effector CD8^+^ T cell frequencies increased dramatically during infection, with peak values ranging from 3.0 × 10^5^ to 2.4 × 10^7^ cells (**Figure 1A**), after scaling their relative abundances to cell quantities, with the population averaging a 1.16 log_10_ increase on day 6 post-infection. Activated memory CD8^+^ T cells began increasing ∼4 days post-infection with individual peaks ranging from 1.5 × 10^6^ to 2.6 × 10^7^ cells, after scaling their relative abundances to cell quantities, with the population averaging a 0.64 log_10_ increase. Most individuals had relatively similar levels of effector and memory CD8^+^ T cells at baseline (**Figure 1B**).

**Figure 1.**
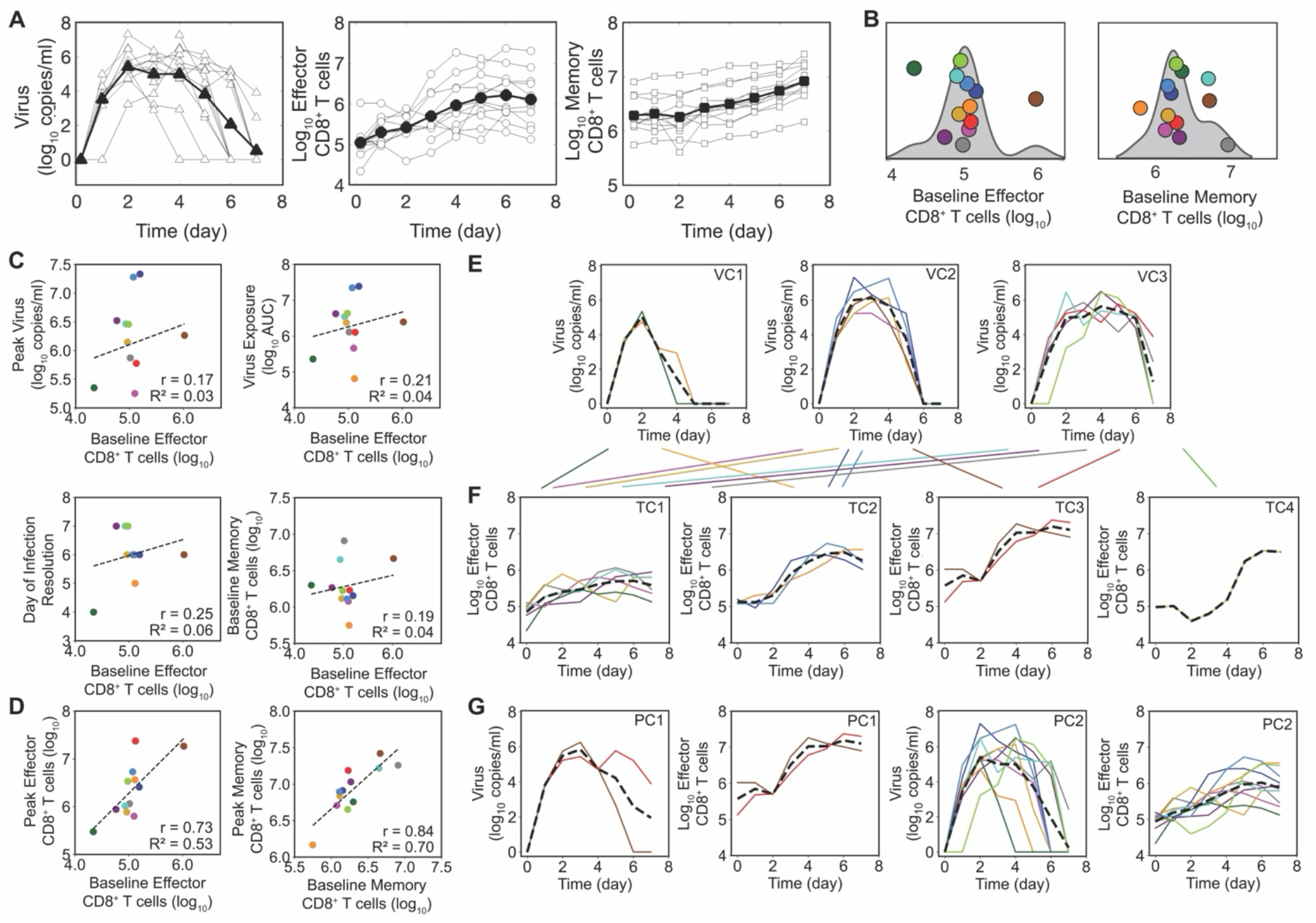
Heterogeneous Viral Shedding and CD8^+^ T cell Response to Experimental Influenza Challenge. (A) Data from experimental influenza A virus challenge in 12 volunteers. Population averages (black markers) and individual trajectories (open markers) are shown for viral shedding (copies/ml) and CD45RA^+^CD38^+^CD8^+^ T cells, including effector [CD27^-^] and memory [CD27^+^] scaled from relative abundances. (B) Baseline CD8^+^ T cell levels were relatively consistent across the cohort.(C) Baseline effector CD8^+^ T cells were not linearly correlated with peak viral load, viral exposure (AUC), time of resolution, or baseline memory CD8^+^ T cell levels. (D) Baseline effector and memory CD8^+^ T cell levels were mildly correlated to their respective peak values. (E-G) Hierarchical clustering performed independently on viral loads (E) and effector CD8^+^ T cells (F) yielded discordant groupings (VC1-3 versus TC1-4), as illustrated by the connecting lines. Clustering based on paired viral load and effector CD8^+^ T cell data identified 2 groups (PC1-2) that lacked strong biological or clinical distinctions (G).

Examining whether baseline effector CD8^+^ T cells levels could be used to predict viral shedding or T cell patterns using linear correlations suggested that baseline effector CD8^+^ T cells were not correlated to virus peak, virus exposure (i.e., area under the curve [AUC]), or day of viral resolution (**Figure 1C**). There were mild correlations between baseline and peak counts for both effector and memory CD8^+^ T cells (r = 0.73 and r = 0.84, respectively; **Figure 1D**). However, for effector CD8^+^ T cells, the correlation was largely skewed by two individuals whose baseline levels were correspondingly above or below most of the cohort, suggesting that baseline levels would likely have minimal predictive power. There were no linear correlations between effector and memory CD8^+^ T cells or virus and memory CD8^+^ T cells (**Figure S2**).

To assess whether measured viral loads and CD8^+^T cell responses could define meaningful infection clusters, we performed hierarchical clustering on each dataset individually and in combination (**Figure 1E-G**). Using only viral loads, the twelve individuals clustered into three groups determined by the day of viral resolution. However, these clusters were not preserved when considering only the effector CD8^+^ T cell data, where a different set of four groups classified by CD8^+^ T cell peak and timing were identified. Clustering based on each individual’s pair of viral loads and CD8^+^ T cells also did not produce meaningful separation, indicating that measured data alone fails to reveal meaningful host-pathogen patterns.

### Viral Infectivity and CD8^+^ T cell Expansion Rates Drive Interindividual Heterogeneity Independent of Exposure Dose

Because T cell data cannot be directly used to predict the course of infection, we sought to establish the predictability of influenza infections and determine how individuals vary in their rates of viral infectivity, replication, and clearance, in addition to CD8^+^ T cell expansion, efficacy, and memory generation to create unique infection trajectories. Fitting a mechanistic mathematical model (Equations (1)-(10)) simultaneously to each individual’s viral load and effector and memory CD8^+^ T cell data accurately captured the observed dynamics, including a biphasic viral decay and heterogeneity in viral loads and CD8^+^ T cell responses between individuals (**Figure 2; Table S1**).

**Figure 2.**
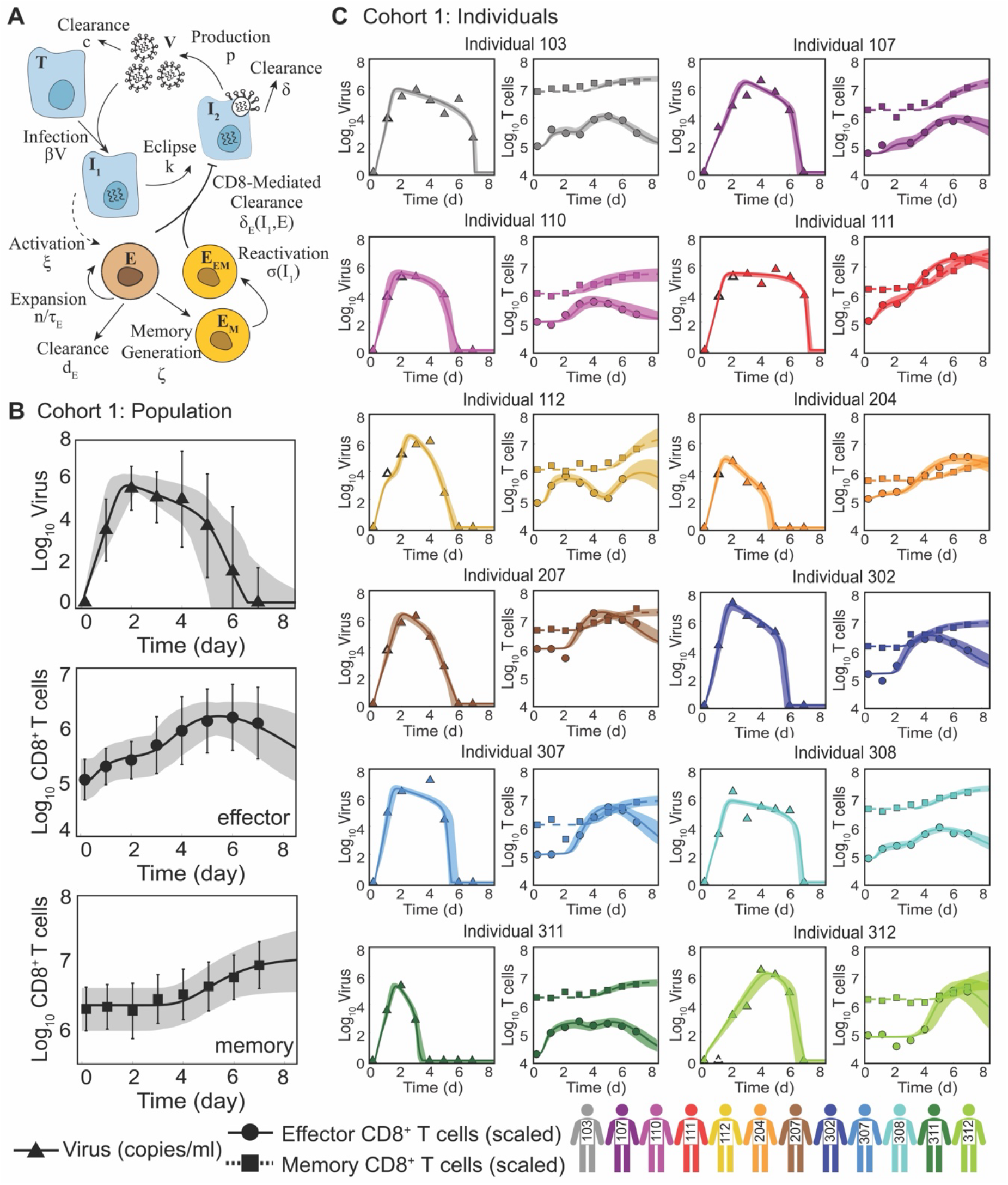
Individualized Mechanistic Modeling of Viral Shedding and CD8^+^ T cell Responses During Human Influenza Infection. (A) Schematic of the mechanistic mathematical model describing human influenza infection in the absence of pre-existing humoral immunity. The model incorporates viral replication, clearance by innate mechanisms, cytotoxic effects of effector CD8^+^ T cells, and the potential reactivation of functional memory CD8^+^ T cells. (B-C) Population-level and individual viral shedding and CD8^+^ T cell responses from Cohort 1, including activated CD45RA^+^CD38^+^ effector (CD27^-^) and memory (CD27^+^) phenotypes shown alongside model fits. Open markers denote imputed viral loads at time points where data were missing. The model reproduces the biphasic viral load decay and captures interindividual variation in the timing and magnitude of viral loads and CD8^+^ T cells.

To separate biological variation from exposure variation, we first accounted for differences in infection size as small variation in viral loads can translate into large differences in the extent of the respiratory tract that becomes infected and in CD8^+^ T cell expansion^34^. Variability in the inoculum delivery likely also contributed, arising from variation in technique, epithelial cell susceptibility, or nasal passage size and structure. The results indicated that infection sizes (T_0_) varied by over two orders of magnitude (2.7 × 10^6^ to 3.4 × 10^8^ cells; **Figure 3A**). This captured the different doses and inoculum volumes used in the challenge study, where volunteers who received the smaller dose (3.5 × 10^6^ TCID_50_ in 250 µl) had lower predicted infection sizes on average compared to volunteers who received the larger dose (7 × 10^6^ TCID_50_ in 500 µl) (**Figure 3A**). Notably, there was variability within each infection group, indicating inherent differential susceptibility amongst the volunteers.

**Figure 3.**
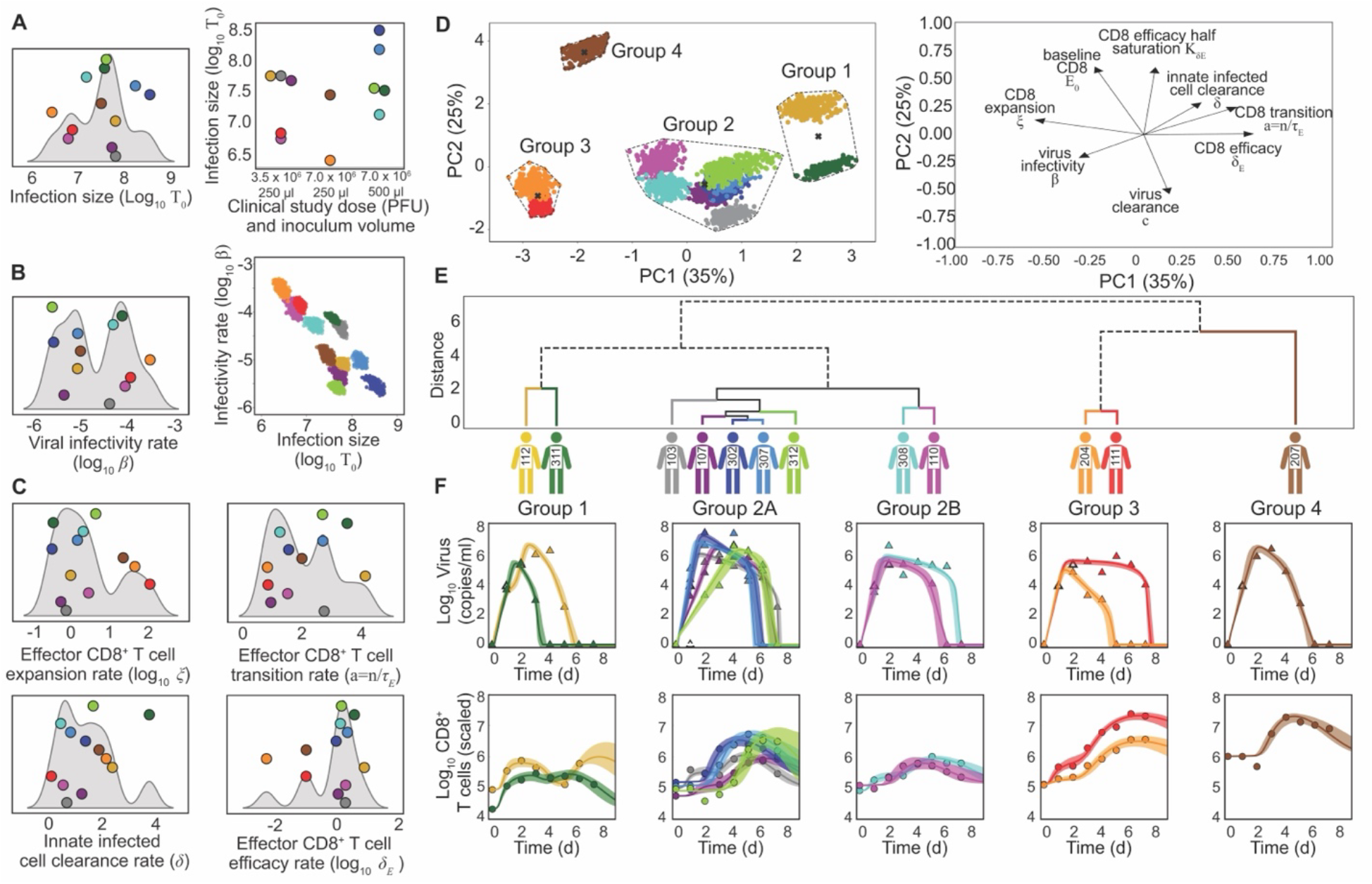
Host-Pathogen Interactions Underlying Individual Heterogeneity in Influenza Virus Infection. (A-C) Interindividual variability in best-fit model parameters for volunteers infected with influenza A virus. Estimated initial infection size (*T*_0_) varied across individuals and aligned with differences in viral dose (A). Viral infectivity rates (*β*) also varied widely and were negatively correlated with infection size across the cohort (B). Variation in parameters governing effector CD8^+^ T cell expansion (*ξ, α* = *n*/*τ*_*E*_) dominated compared to parameters controlling rates of infected-cell clearance by innate mechanisms (*δ*) and CD8^+^ T cells (*δ*_*E*_) (C). Additional parameter distributions are shown in Figure S3. (D) PCA of individualized parameter sets, revealing four distinct groups identified by hierarchical clustering. Loadings illustrate tradeoffs among host-pathogen interaction mechanisms. (E) Dendrogram corresponding to the hierarchical clustering analysis, with branch color and number indicating individual group membership. (F) Corresponding model fits to viral loads (top row; triangles) and scaled effector CD8^+^ T cells (bottom row; circles).

After accounting for differences in dosing, the dominant sources of interindividual heterogeneity were the viral infectivity rate (*β*), which was inversely related to infection size and innate-driven infected cell clearance (*δ*), and effector CD8^+^ T cell expansion rate (*ξ*) (**Figure 3B-C**). In contrast, effector CD8^+^ T cell efficacy (*δ*_*E*_) was largely conserved across the cohort, indicating that much of the apparent variability in T cell measurements reflects scaling with infection size rather than true interindividual differences. This was reflected in a compensatory tradeoff where individuals with lower CD8^+^ T cell efficacy (*δ*_*E*_) exhibited higher expansion rates (*ξ*), yielding comparable functional responses despite differences in per-cell potency (**Figure 3C**; **Figure S3**).

Although this cohort was designed to capture primary influenza infection, two individuals (IDs 112 and 311) exhibited infection dynamics consistent with pre-existing CD8^+^ T cell memory. In these individuals, earlier CD8^+^T cell reactivation led to more rapid infection resolution and was accompanied by higher CD8^+^ T cell efficacy, consistent with functional memory responses despite nominal seronegativity. Differences in innate immune-mediated clearance (*δ*) further contributed to variation in infection resolution, and the broader multivariate heterogeneity emphasizes that similar outcomes can emerge from distinct combinations of host-pathogen interactions.

### Clustering Analysis Revealed Four Distinct Host-Pathogen Patterns

Because interindividual variation spanned multiple processes, we next examined whether multivariate host-pathogen interactions could uncover distinct infection patterns across individuals. To do this, we performed a principal component analysis (PCA) with hierarchical clustering on baseline effector CD8^+^ T cell levels and rates governing viral infection and immunity that determine infection trajectories (**Figure 3D-F**). Infection size (T_0_) was excluded to remove epidemiologic variation, and parameters related to memory reactivation and generation were omitted to focus on host-pathogen interactions active during a primary infection.

The first two principal components accounted for 60% of the variance and separated the twelve individuals in Cohort 1 into four clusters. The analysis naturally grouped individuals with signatures of reinfection, characterized by rapid CD8^+^ T cell expansion with higher efficacy (Cluster 1), and those who received higher viral doses (Cluster 2). Individuals within Cluster 2 further diverged based on their rates of infected cell clearance (δ) and CD8^+^ T cell expansion (ξ).

The remaining two clusters (Clusters 3-4) were characterized by elevated CD8^+^ T cell expansion rates (ξ) and higher viral infectivity (*β*). Tradeoffs between CD8^+^ T cell efficacy (δ_E_) and expansion (ξ) and between viral infectivity rates (*β*) and innate infected cell clearance (δ) were again evident, indicating that stronger innate immunity reduces infectivity and accelerates CD8^+^ T cell differentiation, consistent with antigen-dependent response and the coupling between innate and adaptive immunity.

### Conservation of Host-Pathogen Interactions Across Influenza Challenge Cohorts

To assess whether the host-pathogen interaction patterns identified in Cohort 1 were preserved across studies, we analyzed viral dynamics from a second human influenza challenge cohort infected with a distinct H1N1 influenza strain and dose (1 × 10^5^ TCID_50_ A/Texas/36/91 (H1N1); Cohort 2). Because Cohort 2 reported only viral load measurements and did not include longitudinal immune sampling, we applied a reduced viral dynamics model that implicitly captures CD8^+^ T cell-mediated clearance^54^ (Equations 1-4 with δ(I_2_) = δ_D/_(K_δ_+I_2_); **Figure 4A**; **Figure S4**). The analysis identified higher rates of virus production (*p*) for the influenza A/Texas/36/91 strain compared with influenza A/California/04/2009 (**Table S2**), consistent with strain-specific differences. However, the two cohorts exhibited overlapping infection sizes and host-pathogen interaction rates (**Figure 4B; Figure S4**). As expected given its larger sample size, Cohort 2 displayed greater inter-individual variability, indicating key mechanistic features of infection dynamics are conserved across cohorts despite differences in viral strain and study design.

**Figure 4.**
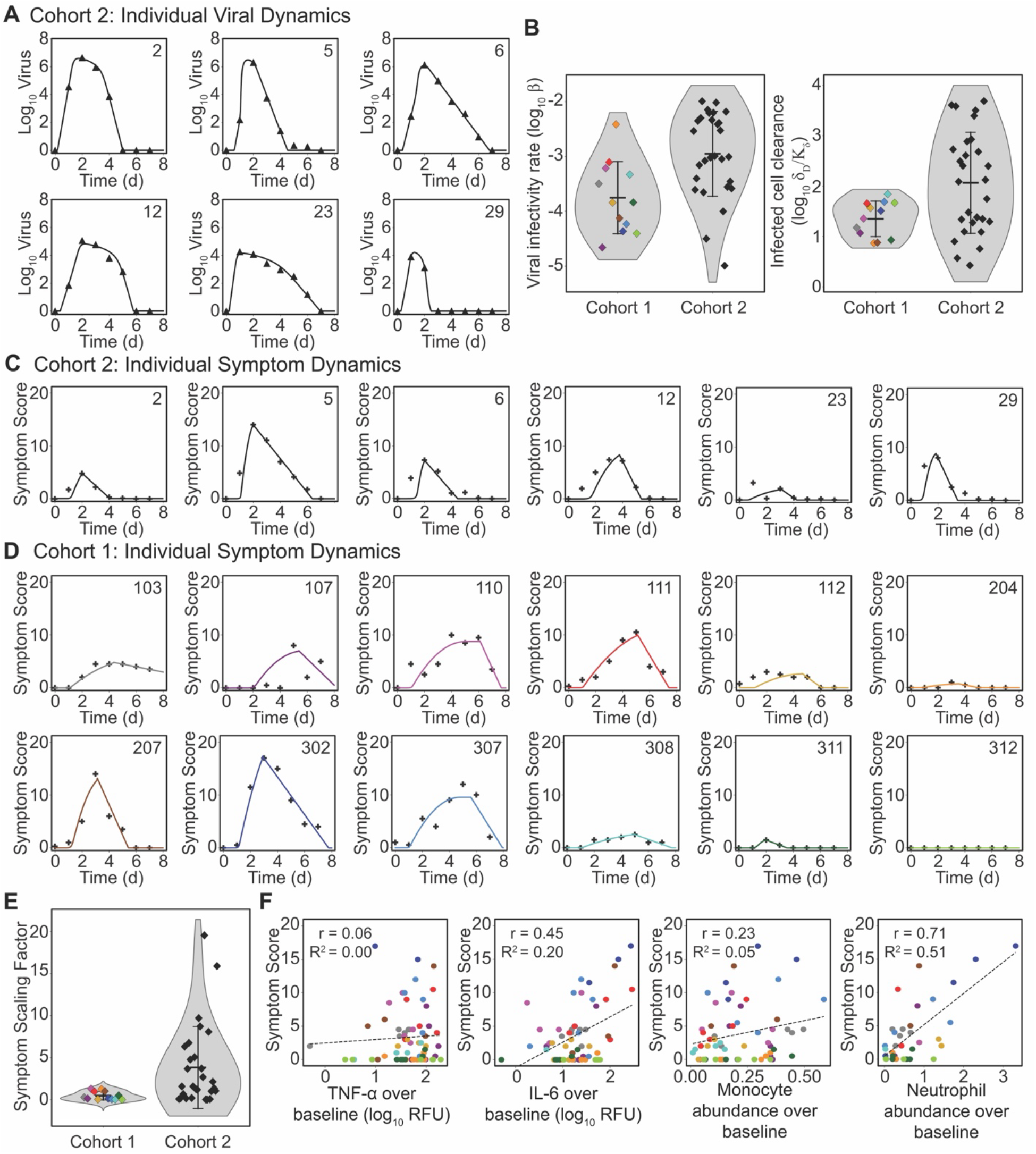
Cross-Cohort Consistency of Host-Pathogen Interaction Dynamics and Symptom Prediction. (A) Model fits to viral load data from representative individuals in Cohort 2; fits for all remaining individuals are shown in Figure S4. (B) Comparison of estimated viral infectivity and infected cell clearance rates between Cohorts 1 and 2, demonstrating conservation of inferred host-pathogen interaction parameters across studies. (C-D) Cumulative area under the curve (CAUC) of model-predicted infected cell dynamics (solid lines) compared with reported symptom scores (markers) for individuals in Cohorts 1 and 2. (E) Comparison of the estimated scaling factor (*α*) linking model-predicted infection burden to symptom scores across cohorts. (F) Linear correlation analysis between symptom scores and circulating inflammatory markers (TNF-α, IL-6) and innate immune cell abundances (monocytes and neutrophils), showing limited correlation.

### Infection Dynamics Predict Symptom Trajectories Despite Individual Subjectivity

We next investigated whether model-predicted infection dynamics could predict the temporal trajectory of symptoms across individuals. Our prior studies demonstrated that the cumulative area under the curve (CAUC) of the predicted infected cell dynamics provides an accurate estimate of infected tissue area and correlates to disease severity^34,58^. Here, we used this same metric to predict human symptoms, which were self-reported and ranged from zero reported symptoms to a peak total score of 17 on a scale of 0 to 51 within Cohort 1, while individuals in Cohort 2 had symptom scores that ranged from 0 to 14 on a scale of 0 to 36 (**Figure 4C-D; Figure S5**).

The results showed good agreement between the CAUC and symptom scores for both Cohorts (**Figure 4C-D**). To convert model-predicted values to symptom scores, we used a scaling factor (α). The average symptom scales were α_1_ = 0.55 (range: 0 to 1.4) for Cohort 1 and α_2_ = 3.8 (range: 0 to 19.6) for Cohort 2 (**Figure 4E**) with substantial overlap between cohorts. These results are consistent with interindividual subjectivity in symptom perception but may instead reflect variability in immune processes related to symptom manifestation, such as cytokines or innate immune activity. To explore this possibility, we examined linear correlations between symptom scores in Cohort 1 and circulating levels of IL-6, TNF-α, and monocyte and neutrophil frequencies (**Figure 4F**). No strong correlations were observed, although symptom scores showed a modest positive association with neutrophil frequencies (r ∼ 0.7).

## Discussion

Identifying how interindividual variation in immunity give rise to heterogeneous infection dynamics and outcomes in human influenza infection is a fundamental challenge to understanding this complicated disease. Although differential baseline immune profiles and CD8^+^ T cell responses have been statistically associated with reduced infection risk, our analyses show that these relationships are more nuanced than correlative studies alone can resolve. This complexity reflects nonlinear host-pathogen relationships that confound direct interpretation of viral shedding and CD8^+^ T cell measurements and increase the likelihood of spurious statistical associations^37^. Through individualized mechanistic modeling, we improve interpretability and predictability and demonstrate that influenza infection trajectories are shaped not by single correlates, but by the interplay of viral dose and interindividual variation in shared viral and host mechanisms, some of which may be clinically unmeasurable. Accounting for these interactions revealed that similar infection trajectories and outcomes can emerge despite biological variation, emphasizing the importance of mechanistic approaches for interpreting immune variability and predicting influenza infection dynamics.

While observable data alone exhibited limited discriminative power to produce stable, biologically meaningful host-pathogen clusters, clustering in mechanistic parameter space revealed that several volunteers grouped together despite variation in estimated rates of viral replication and CD8^+^ T cell responses, indicating that not all heterogeneity is biologically meaningful. Infection trajectories may bifurcate among otherwise similar individuals once key immune or viral processes cross a critical threshold, particularly at higher infectious doses or in the presence of comorbidities. Identifying where these thresholds occur will be critical for understanding and predicting host-pathogen interactions beyond mild infections and for determining the individual-specific points at which T cell responses might shift from protective to immunopathological or confer protection against reinfection. However, new models of innate immunity will be required as viral loads and CD8^+^ T cells do not appear to vary sufficiently to serve as strong indicators of disease severity. In both human and animal studies, macrophages and neutrophils have instead emerged as potential biomarkers of severe influenza^1,7,8,10–12,15–17,19,22^. For Cohort 1, neutrophil frequencies showed the strongest correlation with symptom scores, although they were not required to predict symptom trajectories. Modeling these cells will be important to better predict divergent outcomes for influenza.

The infection variability resulting from differences in viral doses and infectivity rates and cross-cohort differences in viral production rates emphasizes the challenge in predicting influenza infections when initial conditions and viral strain are unknown. Beyond transmission route or, in experimental challenges, infection technique, several factors may contribute to smaller initial viral seedings, including enhanced phagocytosis by tissue-resident macrophages and dendritic cells, as well as genetic or functional variation in epithelial cells. This may align with our findings of high interindividual viral infectivity rates and with studies that statistically associate higher frequencies of NK cells, monocytes, and *γδ* T cells in the blood with increased susceptibility and disease^1–3,8,11,12,15,17,22,23,59^, although we caution against overinterpreting such correlations as most studies lack time resolution to identify early deviation points.

Most individuals exhibited similar CD8^+^ T cell efficacy but differed substantially in their rates of CD8^+^ T cell expansion. This suggests that among otherwise healthy individuals, infection outcomes are shaped less by qualitative differences in cytotoxic function than by quantitative differences in the dynamics of immune deployment relative to infection burden. This interpretation is consistent with our prior murine influenza modeling and T cell depletion experiments, which showed that reducing CD8^+^ T cells by more than 2 log_10_ did not alter the rate of virus clearance^34^, emphasizing that T cell counts are best interpreted with the help of mechanistic models and in the context of viral loads or other infection characteristics. In those who deviated from this pattern, T cell efficacy was negatively related to expansion rate, suggesting that larger numbers of T cells compensate for reduced per-cell function. In contrast, innate immune clearance of infected cells, which largely determines the slope of viral decline after the peak^60^, showed greater variability, likely leading to differences in the timing of T cell responses influenced by early dendritic cell activity.

Our analysis identified two individuals exhibiting evidence of functional memory CD8^+^ T cell responses, which was characterized by early increases in CD8^+^ T cells, despite being seronegative. Smaller early increases were also observed in several other individuals but did not reach levels consistent with functionally relevant reactivation. Importantly, inferred host-pathogen interaction parameters fell within similar ranges across all individuals, indicating that the mechanistic framework reliably predicts both primary infection and infections occurring in the presence of partial cellular memory. Whether the rapid CD8^+^ T cell responses observed in these individuals reflects immunity to the challenge strain or cross-reactive memory from prior influenza exposure remains unclear and may explain why viral replication and shedding still occurred. Consistent with this interpretation, a prior human challenge study found that nearly all individuals were reinfected one year after initial exposure^41^. Greater cohort size and immunological resolution would enable more precise quantification of heterogeneity in CD8^+^ T cell reactivation and assessment of how baseline memory influences infection trajectories. Reinfection and symptom severity likely vary with waning immunity, exposure history, and strain differences, yet remain interpretable and predictable within our mechanistic modeling framework.

Predicting symptom dynamics is central to understanding influenza severity. Our mechanistic model links symptom manifestation to the cumulative number of infected cells, consistent with prior mouse studies showing that symptom severity reflects tissue damage^34,58^. It also reproduces earlier findings that small viral load differences can produce large variations in symptom severity and accounts for individuals who shed virus without noticeable symptoms^34^. The relationship between infection dynamics and symptom expression suggests interindividual differences in how tissue-level immunity translates to clinical symptoms. This modeling metric underestimates early symptoms in some individuals, potentially reflecting systemic responses like fever rather than respiratory symptoms like coughing^61,62^. Future models incorporating validated representations of innate immune responses should clarify their contribution to protection from symptomatic influenza. Overall, these results highlight the utility of mechanistic modeling for predicting individual outcomes beyond what peripheral immune measures can explain.

Our results collectively highlight the importance of accounting for exposure dose and host immune responses when interpreting viral shedding dynamics. Our analyses were limited by the original datasets, which lacked time-resolved measurements of absolute cell counts, constraining the resolution of infection dynamics. Viral loads alone provide minimal insight into innate immunity or other factors that shape protection and disease severity. In natural infections, where multiple influenza strains circulate and evolve, heterogeneity in dose and viral replication further complicates interpretation. Detailed immune measurements combined with mechanistic modeling are therefore essential to disentangle these complex dynamics.

These findings demonstrate the value of individualized mechanistic modeling for predicting influenza infection dynamics and disease severity by moving beyond single correlates to capture multivariate virus-host interactions. This individualized parameterization represents a first step toward mechanistic digital twins of human influenza infection, person-specific computational models that causally link viral replication, immune response, and symptom dynamics at the level of the individual, with broad implications for personalized risk prediction, vaccine evaluation, and the design of targeted interventions. By focusing on how interindividual heterogeneity manifests in infection trajectories, this framework accommodates substantial variation in human immunity while still yielding interpretable and predictive insights. Although important questions remain regarding the explicit contributions of innate, humoral, and trained immunity, our results identify important interindividual variation and show that mechanistic inference substantially improves the ability to forecast heterogeneous infection dynamics across individuals and cohorts.

## Data Availability

All data produced in the present work are contained in the manuscript

## Acknowledgments

This work was supported by the NIH NIAID under award numbers R01AI170115 and R01AI139088 and, in part thanks to the Canada Research Chair in Computational Immunology with funding from the Canada Research Chairs Program. We thank Jonathan McCullers for helpful comments, and Andrew Gienapp for technical editing.

