## Supplemental Figures for "Individualized Mechanistic Modeling Reveals Viral Infectivity and CD8^+^ T cell Expansion as Drivers of Heterogeneous Influenza Dynamics"

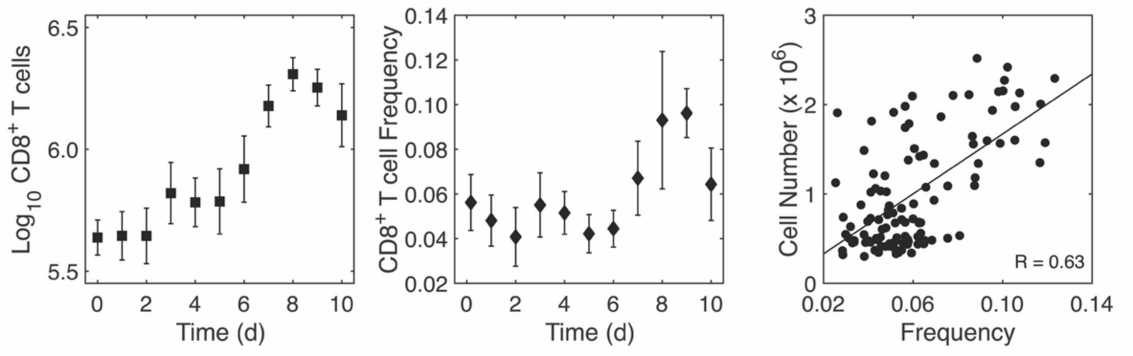


**Figure S1. Relation Between CD8$^+$ T cell Number and Frequencies in Murine Influenza Virus Infection.** Total CD8$^+$ T cell counts and frequencies in BALB/c mice infected with influenza A/PR8. Cell numbers were derived by scaling CD8$^+$ T cell frequencies to the total lung cell count. Frequencies were calculated as the number of CD8$^+$ T cells per $10^5$ cells analyzed by flow cytometry. The increase beginning around days 5-6 coincides with the period when T cells predominate during infection. Cell numbers and frequencies were only moderately correlated.

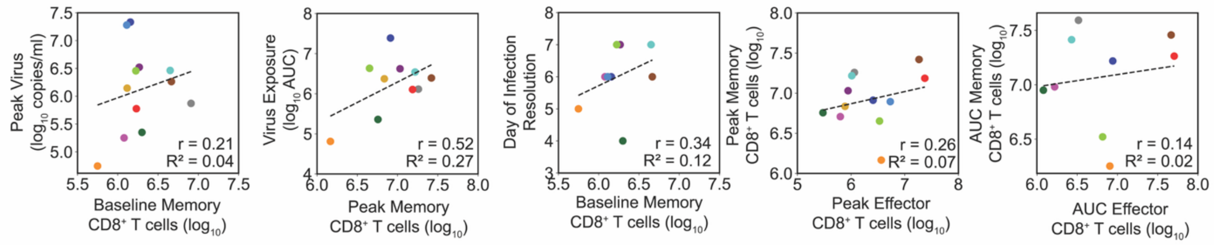


**Figure S2. Additional Linear Correlation Analysis Between Virus and CD8⁺ T cells.** Baseline and peak memory CD8⁺ T cells were not correlated with virus peak, viral exposure (area under the curve; AUC), day of resolution, or peak effector CD8⁺ T cells. The AUC of effector CD8⁺ T cells was also not correlated with the AUC of memory CD8⁺ T cells.

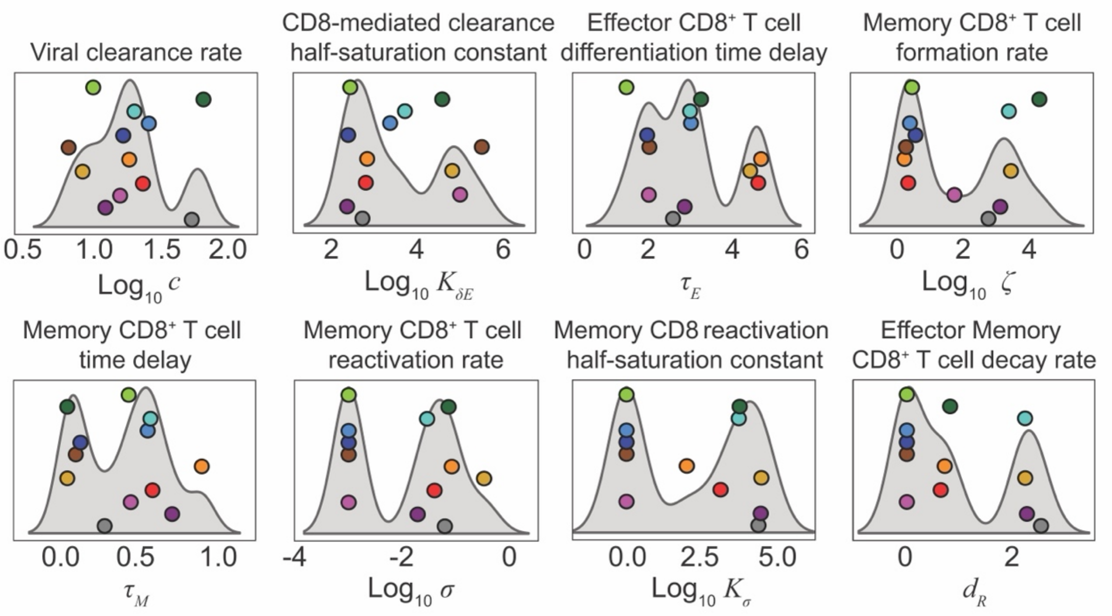


**Figure S3. Additional Host-Pathogen Interaction Parameters Underlying Heterogeneous Viral Shedding and CD8⁺ T cell Dynamics during Influenza Virus Infection.** Interindividual variability in best-fit mechanistic model parameters for volunteers infected with influenza A virus in Cohort 1 that are not shown in the main text. Complementary parameter distributions in Figure 3 summarize the primary parameters discussed in the Results.

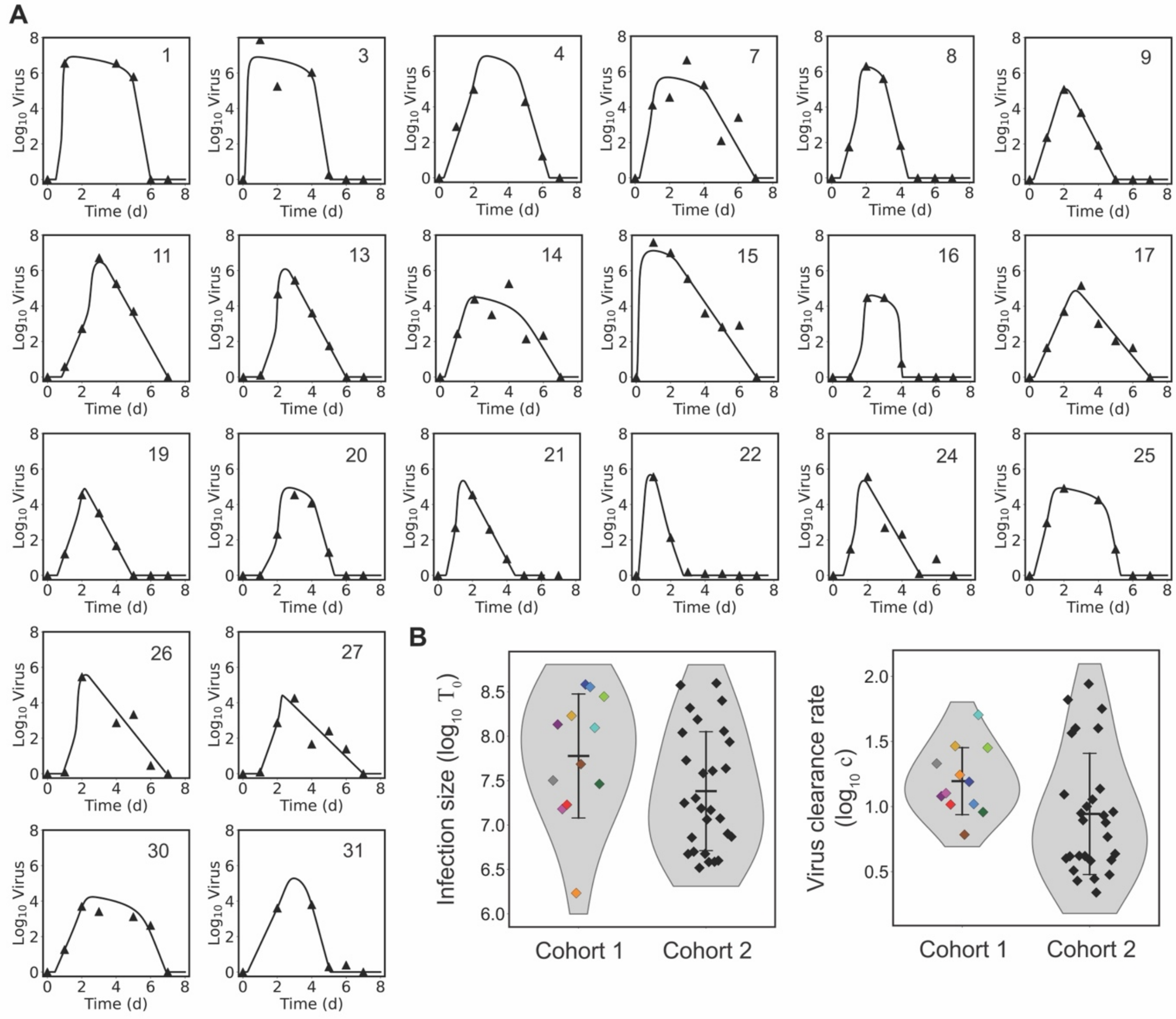


**Figure S4. Additional Individualized Mechanistic Modeling of Viral Shedding Dynamics in Cohort 2.** (A) Fits of a reduced viral kinetic model (Equations 1-4 with $\delta(I_2) = \delta_D/(K_\delta + I_2)$) to viral shedding data from individuals in Cohort 2 not shown in the main text. (B) Comparison of the estimated infection sizes and rates of virus clearance between Cohorts 1 and 2, demonstrating overlap in inferred host-pathogen interactions despite differences in viral strain and study design.

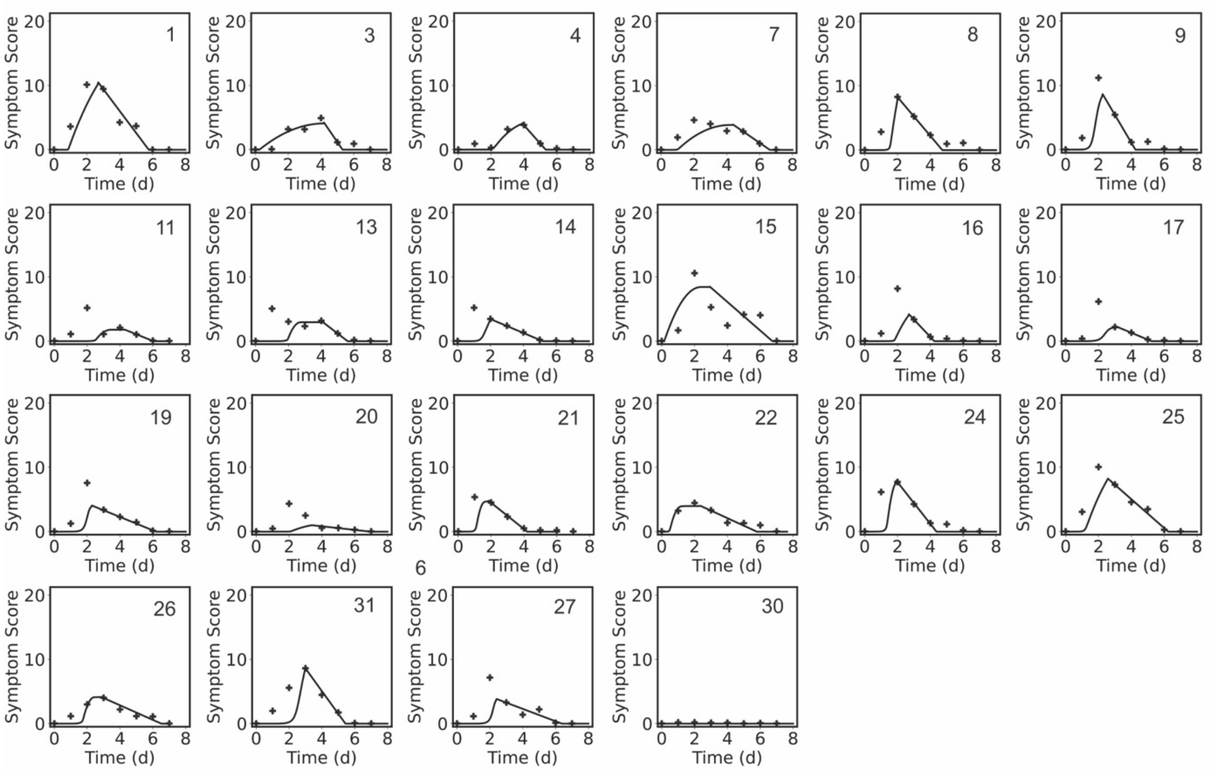


**Figure S5. Additional Model-Predicted Symptom Dynamics in Cohort 2.** Cumulative area under the curve (CAUC) of model-predicted infected cell dynamics (solid lines) compared with reported symptom scores (markers) from individuals in Cohort 2 from not shown in the main text.
